# Previous COVID-19 infection but not Long-COVID is associated with increased adverse events following BNT162b2/Pfizer vaccination

**DOI:** 10.1101/2021.04.15.21252192

**Authors:** Rachael K. Raw, Clive Kelly, Jon Rees, Caroline Wroe, David R. Chadwick

**Affiliations:** School of Medicine and Health, Newcastle University, Newcastle upon Tyne, UK; The Department of Rheumatology, James Cook University Hospital, Middlesbrough, UK; The School of Psychology, University of Sunderland, UK; The Department of Nephrology, James Cook University Hospital, Middlesbrough, UK; Centre for Clinical Infection, James Cook University Hospital, Middlesbrough, UK

## Abstract

**Importance:** Understanding Adverse Events (AEs) associated with SARS-CoV-2 vaccination has public health implications, especially with regards to vaccine hesitancy.

**Objective:** To establish whether individuals with prior history of COVID-19 were more likely to experience AEs after BNT162b2/Pfizer vaccination, than those without previous COVID-19, and whether COVID-19-vaccination interval influenced AE severity.

**Design:** An observational study explored AEs after vaccination. Participants were invited to complete an electronic survey, capturing self-reported COVID-19 symptoms, PCR/antibody results, and AEs following first dose of BNT162b2/Pfizer vaccine. In a subset where PCR/antibody results could be verified, a sensitivity analysis was conducted.

**Setting:** Three North-East England hospital Trusts in the United Kingdom.

**Participants:** Healthcare workers formed an opportunistic sample – 265 of 974 reported prior positive SARS-CoV-2 PCR and/or antibody.

**Exposure:** All participants had received their first dose of BNT162b2/Pfizer vaccine.

**Main Outcomes and Measures:** Nature, severity, duration, and onset of self-reported AEs (reported via a modified version of the FDA Toxicity Grading Scale for vaccine-associated AEs), was compared between those with and without a prior history of COVID-19, using 2-way ANCOVA and logistic regression. Effects of age, gender, illness-vaccine interval, and ongoing symptoms (‘Long-COVID’) on AEs, were also explored.

**Results:** Of 974 respondents (81% female, mean age 48), 265 (27%) reported previous COVID-19 infection. Within this group (symptoms median 8.9 months pre-vaccination), 30 (11%) complained of Long-COVID. The proportion reporting one moderate/severe symptom was higher in the previous COVID-19 group (56% v 47%, OR=1.5 [95%CI, 1.1–2.0], p=.009), with fever, fatigue, myalgia-arthralgia and lymphadenopathy significantly more common. There was no significant relationship between illness-vaccine interval and symptom composite score (r_s_=0.09, p=.44). Long-COVID was not associated with worse AEs in comparison to the group without previous COVID-19. In the smaller sensitivity analysis cohort (412 people) similar findings were obtained although only myalgia and arthralgia remained significant.

**Conclusions and Relevance:** Prior COVID-19 infection but not ongoing Long-COVID symptoms were associated with an increase in the risk of self-reported adverse events following BNT162b2/Pfizer vaccination. COVID-19 illness-vaccination interval did not significantly influence AEs. This data can support education around vaccine-associated AEs and, through improved understanding, help to combat vaccine hesitancy.

**Key Points:** *Question:* Does previous COVID-19 infection or ‘Long-COVID’ increase the frequency of Adverse Events (AEs) following first dose of BNT162b2/Pfizer vaccination?

*Findings:* In a survey-based observational study, healthcare workers in the United Kingdom reported AEs experienced after their first dose of BNT162b2/Pfizer vaccine. Prior COVID-19 infection, but not Long-COVID, were associated with increased risk of self-reported AEs including lymphadenopathy post-vaccination. Duration since COVID-19 infection did not affect severity of AEs.

*Meaning:* Our study can inform education and understanding of AEs associated with COVID-19 vaccination and help to combat vaccine hesitancy.

## Introduction

The BNT162b2/Pfizer and mRNA-1273/Moderna COVID-19 vaccines^1,2^ were recently approved for use in the UK, with the former widely used amongst priority groups. While safety profiles were deemed acceptable (following phase 3 trials), participants with previous COVID-19 infection were excluded. Recent evidence suggests mRNA vaccines may cause more Adverse Events (AEs) in those with a history of COVID-19.^3-5^ A small study found that AEs reported after the first dose of mRNA vaccine in seropositive individuals, were greater than in those with no prior COVID-19.^3^ The ‘ZOE COVID-19 Symptom Study’ also observed similar outcomes via a self-reporting app.^4^ Most recently in a larger study, 532 out of 2002 participants with prior COVID-19 reported increased (mostly systemic) AEs after either an mRNA or vector-based (AZD1222/AstraZeneca) vaccine.^5^

These preliminary studies suggest a need for further investigation into the effect of prior COVID-19 history on vaccine-related AEs. Consideration of whether time between previous infection and vaccination administration or the presence of ‘Long-COVID’^6-8^ can predict AEs, is also warranted. This information is important, as it could assist in identifying individuals who are more likely to experience side effects to COVID-19 vaccines. Furthermore, there are public health implications with regards to vaccine hesitancy, which is somewhat driven by fear of AEs.^9-11^ As part of a longitudinal observational study of COVID-19 in healthcare workers in North-East England, we evaluated AEs following first doses of BNT162b2/Pfizer vaccine, with particular reference to previous COVID-19 and Long-COVID.

## Method

National Health Service (NHS) workers (employed by 3 North-East Trusts in the UK) completed an electronic survey on AEs following COVID-19 vaccination. The survey captured self-reported COVID-19 symptoms, PCR/antibody results, and AEs following the first dose. The FDA Toxicity Grading Scale^12^ (with simplified language) was modified allowing participants to self-report AEs for severity (mild/moderate/severe/very severe), duration (≤24 hours/>24 hours) and onset (≤24 hours/>24 hours); lymphadenopathy was included as an additional symptom.

A composite score for symptom nature and severity was calculated, to provide an overall estimate of AE-related morbidity, for the former by adding number of moderate/severe symptoms, and the latter by multiplying this by symptom duration. Individual and composite AE scores were compared between those with and without a prior history of COVID-19, as indicated by self-reported prior positive antibody and/or PCR result. Long-COVID was defined as symptoms persisting >2 months to vaccination. Effects of age, gender and time between past infection to vaccination were also considered.

Respondents who had permitted laboratory results to be accessed (SARS-CoV-2 PCR and antibody), formed a subgroup for sensitivity analysis. Statistical analysis was carried out using JASP v0.14.1.0. Composite scores were compared using 2-way ANCOVA. Multivariable logistic regressions were performed to identify the relationship between COVID-19 status and the presence of moderate/severe symptoms in each category, and the Bonferroni correction applied to the resulting significance and confidence intervals. The study was approved by Cambridge East Research Ethics Committee.

## Results

Of 974 healthcare workers (aged 19-72-years) responding to the survey and providing complete data for analysis, 265 (27%) participants (84% female, mean age 48.9) reported a prior positive PCR and/or antibody result, and 709 (80% female, mean age 47.0) had no COVID-19 history. Within the previous COVID-19 group (symptoms median 8.9 months before vaccination), 30 (83% female, mean age 48.8) complained of Long-COVID (median duration 9.3 months, range 2.8–10.4).

Figure 1A shows frequencies of each symptom by COVID-19 status. The proportion of participants reporting at least one moderate-to-severe symptom was higher in the previous COVID-19 group (56% v 47%, OR=1.5 [95%CI, 1.1–2.0], p=.009). Symptom onset was mostly within 24 hours (75%) with no onset >48 hours. Number and total duration of reported symptoms was greater in women (1.24 (1.67) v 0.84 (1.46) symptoms, d=0.25 [0.09–0.42], p=.002; 2.10 (2.99) v 1.39 (2.54) symptom-days, d=0.22 [0.09–0.42], p=.001) and significantly decreased with age (symptoms: r_s_=-0.25, p<.001; symptom-days: r_s_=-0.24, p<.001). After controlling for age and sex, higher symptom number (1.61 (2.26) v 0.89 (2.02) symptoms, d=0.34 [0.20-0.49], p<.001) and severity (2.7 (6.65) v 1.5 (2.21) symptom-days, d=0.41 [0.27-0.55], p<.001) were significantly associated with reporting previous COVID-19.

**Figure 1.**
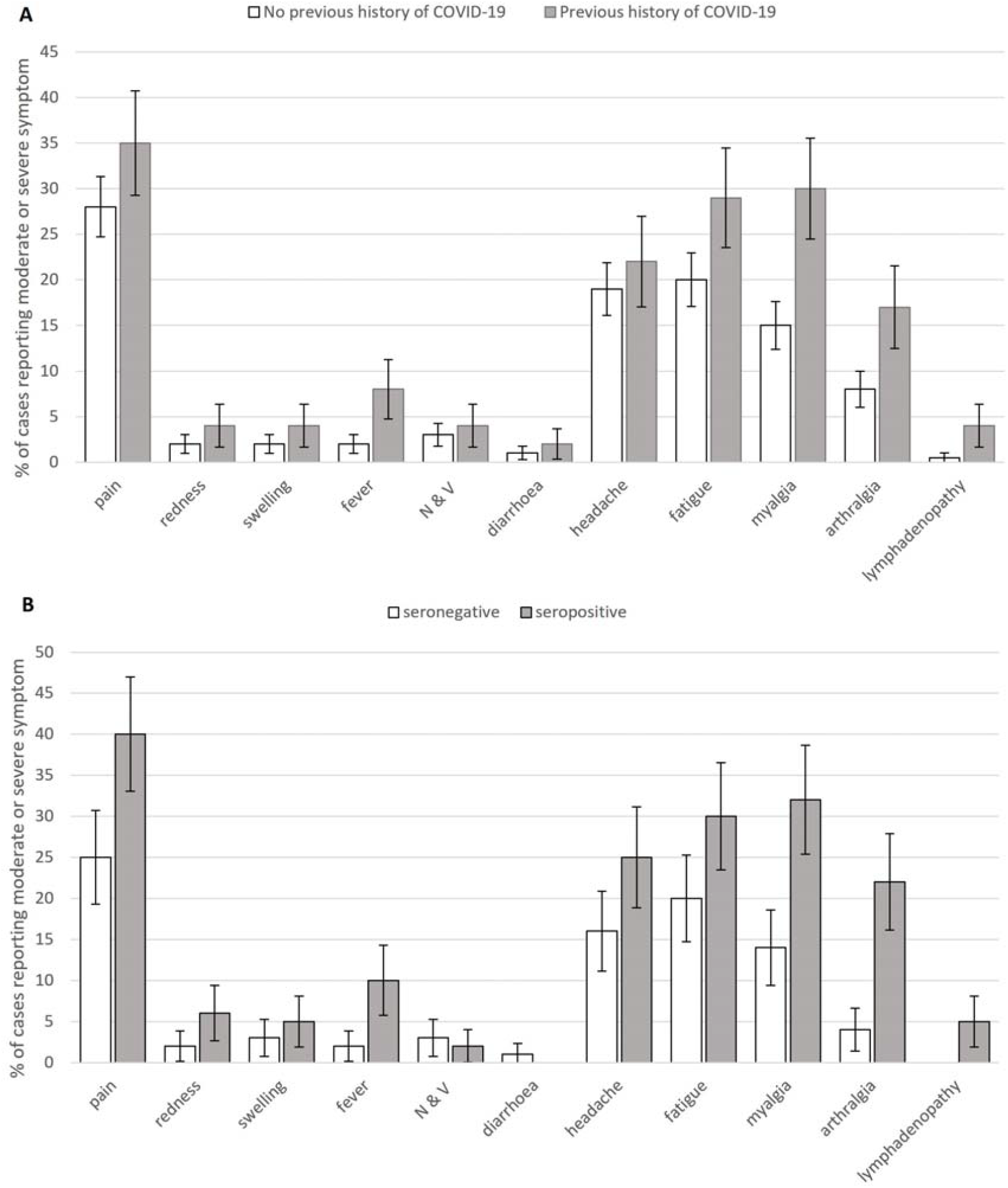
Moderate and Severe Symptoms by COVID-19 Status: Percentage of cases reporting moderate or severe symptoms (95% CI) in those with and without a history of COVID-19. N & V: nausea and vomiting. Upper panel (A): entire cohort; lower panel (B): sensitivity analysis subset

Logistic regressions (Table 1) controlling for age and sex showed five systemic symptoms were significantly associated with previous COVID-19 status: fever, fatigue, myalgia, arthralgia and lymphadenopathy. Arthralgia was regularly co-reported with myalgia (87 cases) but rarely alone and was not independently associated (OR 1.4 [95%CI 0.86–2.37], p=0.49) with COVID-19 exposure once myalgia was controlled for. Neither local nor gastrointestinal symptoms were significantly associated with previous COVID-19 history.

**Table 1.** Results of Logistic Regression Analyses: Logistic regressions showing those symptoms significantly predicted by previous history of COVID-19 after controlling for differences in age and gender and with p values and confidence intervals corrected (Bonferroni) for multiple comparisons.

|  | Whole cohort |  | Sensitivity Analysis Subset |  |
| --- | --- | --- | --- | --- |
|  | Odds Ratio (95% C.I.) | p | Odds Ratio (95% C.I.) | p |
| Fever | 2.87 (1.10 – 7.51) | .044 | 5.68 (0.69 – 46.65) | .32 |
| Fatigue | 1.78 (1.12 – 2.84) | .011 | 2.17 (0.85– 5.54) | .31 |
| Myalgia | 2.34 (1.44 – 3.88) | <.001 | 3.18 (1.16 – 8.69) | .02 |
| Arthralgia | 2.25 (1.23 – 4.12) | .004 | 7.06 (2.05 – 36.91) | .01 |
| Lymphadenopathy | 5.18 (1.19 – 22.63) | .033 | **** | **** |
| Local Pain | 1.55 (0.99 – 2.40) | .09 | 2.28 (0.96 – 5.43) | .11 |
| Local Redness | 2.93 (0.84 – 10.20) | .24 | 3.92 (0.43 – 35.79) | >.99 |
| Local Swelling | 2.0 (0.64 – 6.27) | .14 | 2.1 (0.29 – 15.33) | >.99 |
| N & V | 1.47 (0.48 – 4.42) | >.99 | 0.72 (0.05 – 8.81) | >.99 |
| Diarrhoea | 2.35 (0.30 – 18.25) | >.99 | **** | **** |
| Headache | 1.31 (0.80 – 2.15) | >.99 | 1.78 (0.65 – 4.83) | >.99 |

\*\*\*\* No model could be calculated due to absence of cases in this cohort. In all cases age and gender were included in the null model as nuisance variables. Adjusted P values and adjusted confidence intervals corrected (Bonferroni) for 11 outcomes in each case.

Symptom number and duration was not significantly higher in those with Long-COVID after accounting for gender and age effects and no individual symptom was significantly associated with this condition. Importantly, among those with prior COVID-19, there was no significant relationship between illness-vaccine time interval and either composite score (r_s_=0.09 p=.44 for symptoms; r_s_=0.10, p=.42 for symptom–days) nor any difference in mean time interval based on presence of any of the symptoms (all p>0.05).

For the sensitivity analysis, 412 participants had verified PCR/antibody results. Of this subgroup, 228 (55%) were PCR/antibody negative (80% female, mean (SD) age 47.0 [11.1]) and 184 (45%) PCR or antibody positive (91% female, mean (SD) age 47.3 [11.5]). Nine (5%) complained of Long-COVID (range 2.8–10.4 months). The pattern of results was broadly replicated in this subgroup analysis (Figure 1B), with more previous-COVID-19 individuals reporting at least one moderate symptom (63% v 43%, OR=2.2 [1.2–4.0], p=.006) and previous-COVID-19 being associated with higher symptom number (1.81 (3.09) v 0.85 (4.12) symptoms, d=0.25 [0.05–0.44] p=.012) and severity (3.0 (8.3) v 1.5 (5.6) symptom days d=0.2 [95% CI 0.02–0.41], p=.0350). Only myalgia and arthralgia remain as significant outcomes once multiple comparisons were controlled for though pattern of outcomes remains similar.

## Discussion

This study of healthcare workers demonstrated that prior COVID-19 infection, but not Long-COVID, is associated with increased risk of AEs including lymphadenopathy following BNT162b2/Pfizer vaccination, although there was no relationship with duration since COVID-19 illness. Women and younger individuals were also more likely to experience vaccine-related AEs. Our findings add to other reports supporting wider understanding of AEs following COVID-19 vaccination.^3-5^ Importantly, given the hesitancy surrounding COVID-19 vaccination,^9-11^ our findings may help inform those with previous COVID-19, including Long-COVID, of increased susceptibility to certain AEs. Our study also adds weight to the question of whether a second dose of mRNA vaccine is necessary in those with previous COVID-19, assuming effective immunity is established after the first dose.^3,14^ This is relevant, given that another study has suggested worse AEs following the second dose.^5^

Our study has several limitations. Firstly, some non-responder bias^13^ is likely, with 27% of participants reporting previous COVID-19. This is slightly higher than in UK healthcare workers.^15^ Nevertheless, the sample was broadly representative of UK healthcare employees and likely generalizable. Secondly, information on AEs was gathered via self-reported questionnaires, and hence subjective. Thirdly, PCR and antibody results were self-reported. We addressed this via a sensitivity analysis on a subset of participants with laboratory data available, which mostly confirmed the findings in the entire sample. Finally, the numbers with Long-COVID were relative small for comparison.

In conclusion, this large study shows an association of previous COVID-19 with increased AEs and will help those with previous COVID-19 infection understand better what to expect following vaccination.

## Data Availability

As no consent was obtained from participants to allow data to made generally available, this data is restricted.

## Author Contributions

DRC/CK/RKR conceived the study and DRC is chief investigator of CHOIS. RKR acted as site principal investigator. DRC/RKR/CW contributed to the study protocol, design, and data collection. JR/RKR/DRC did the statistical analysis. RKR/JR/DRC prepared the manuscript. All authors critically reviewed and approved the final version.

## Acknowledgements

We would like to thank the CHOIS research team, John Rouse and the North East and North Cumbria NIHR for assistance with the survey. Funding for the CHOIS study is from the North East and North Cumbria Academic Health Sciences Network (AHSN) and Siemens Healthcare Ltd, who provided assays – but had no input into the study design.

